# Extending Inherited Metabolic Disorder diagnostics with Biomarker Interaction Visualizations

**DOI:** 10.1101/2022.01.31.21265847

**Authors:** Denise N. Slenter, Irene M.G.M. Hemel, Chris T. Evelo, Jörgen Bierau, Egon L. Willighagen, Laura K.M. Steinbusch

## Abstract

**Background:** Inherited Metabolic Disorders (IMDs) are rare diseases where one impaired protein leads to a cascade of changes in the adjacent chemical conversions. IMDs often present with non-specific symptoms, a lack of a clear genotype-phenotype correlation, and *de novo* mutations, complicating diagnosis. Furthermore, products of one metabolic conversion can be the substrate of another pathway obscuring biomarker identification and causing overlapping biomarkers for different disorders. Visualization of the connections between metabolic biomarkers and the enzymes involved might aid in the diagnostic process. The goal of this study was to provide a proof-of-concept framework for integrating knowledge of metabolic interactions with real-life patient data before scaling up this approach. This framework was tested on two groups of well-studied and related metabolic pathways (the urea cycle and pyrimidine de-novo synthesis). The lessons learned from our approach will help to scale up the framework and support the diagnosis of other less understood IMDs.

**Methods:** Our framework integrates literature and expert knowledge into machine-readable pathway models, including relevant urine biomarkers and their interactions. The clinical data of 16 previously diagnosed patients with various pyrimidine and urea cycle disorders were visualized on the top 3 relevant pathways. Two expert laboratory scientists evaluated the resulting visualizations to derive a diagnosis.

**Results:** The proof-of-concept platform resulted in varying numbers of relevant biomarkers (five to 48), pathways and pathway interactions for each patient. The two experts reached the same conclusions for all samples with our proposed framework as with the current metabolic diagnostic pipeline. For nine patient samples the diagnosis was made without knowledge about clinical symptoms or sex. For the remaining seven cases, four interpretations pointed in the direction of a subset of disorders, while three cases were found to be undiagnosable with the available data. Diagnosing these patients would require additional testing besides biochemical analysis.

**Conclusion:** The presented framework shows how metabolic interaction knowledge can be integrated with clinical data in one visualization, which can be relevant for future analysis of difficult patient cases and untargeted metabolomics data. Several challenges were identified during the development of this framework, which should be resolved before this approach can be scaled up and implemented to support the diagnosis of other (less understood) IMDs. The framework could be extended with other OMICS data (e.g. genomics, transcriptomics), phenotypic data, as well as linked to other knowledge captured as Linked Open Data.

## Background

Many enzymes are critically involved in the synthesis, degradation, and transport of molecules in metabolic processes [1]. Malfunctioning of any of these enzymes often results in a lack of or (potentially) toxic levels of metabolites, as well as affecting other (downstream) pathways [2]. Figure 1 presents a schematic of the disturbed biochemical reactions based on one impaired protein, leading to an altered phenotype. These disorders are classified as Inherited Metabolic Disorders (IMDs) or Inborn Errors of Metabolism [3]. A timely and accurate diagnosis of IMDs, currently based on both symptoms and biomarkers measured in various bodily fluids, is required to initiate therapies, which are sparsely available [4]. The current diagnostic process starts with a metabolic pediatrician, who based on the phenotype of a patient can request biochemical analysis on a patient sample (e.g. blood, urine). After the sample has been collected and processed, several types of analysis can be performed (e.g targeted metabolite assays, Whole Exome Sequencing (WES)), which all require data processing and interpretation. The processed data is often linked to existing database knowledge to arrive at a diagnosis. Methods to detect genetic variants (WES) are useful for the diagnosis of specific classes of IMDs where few or no specific metabolic biomarkers exist (e.g. mitochondrial disorders). This technique has been found less sensitive and specific as compared to metabolic measurements in newborn screening [5]. Furthermore, genetic profiles of patients can also contain variants of uncertain significance (Figure 1); these variants can only be classified as (likely) pathogenic when genomic, transcriptomic, proteomic, metabolomic, and/or fluxomic data are integrated through pathway or network analysis [6–8]. Targeted metabolite assays on the other hand are a valuable tool to pinpoint which metabolic processes are disturbed, if the biomarkers for a disorder are known. These altered metabolites are used in newborn screening through dried blood spot analysis.

**Figure 1:**
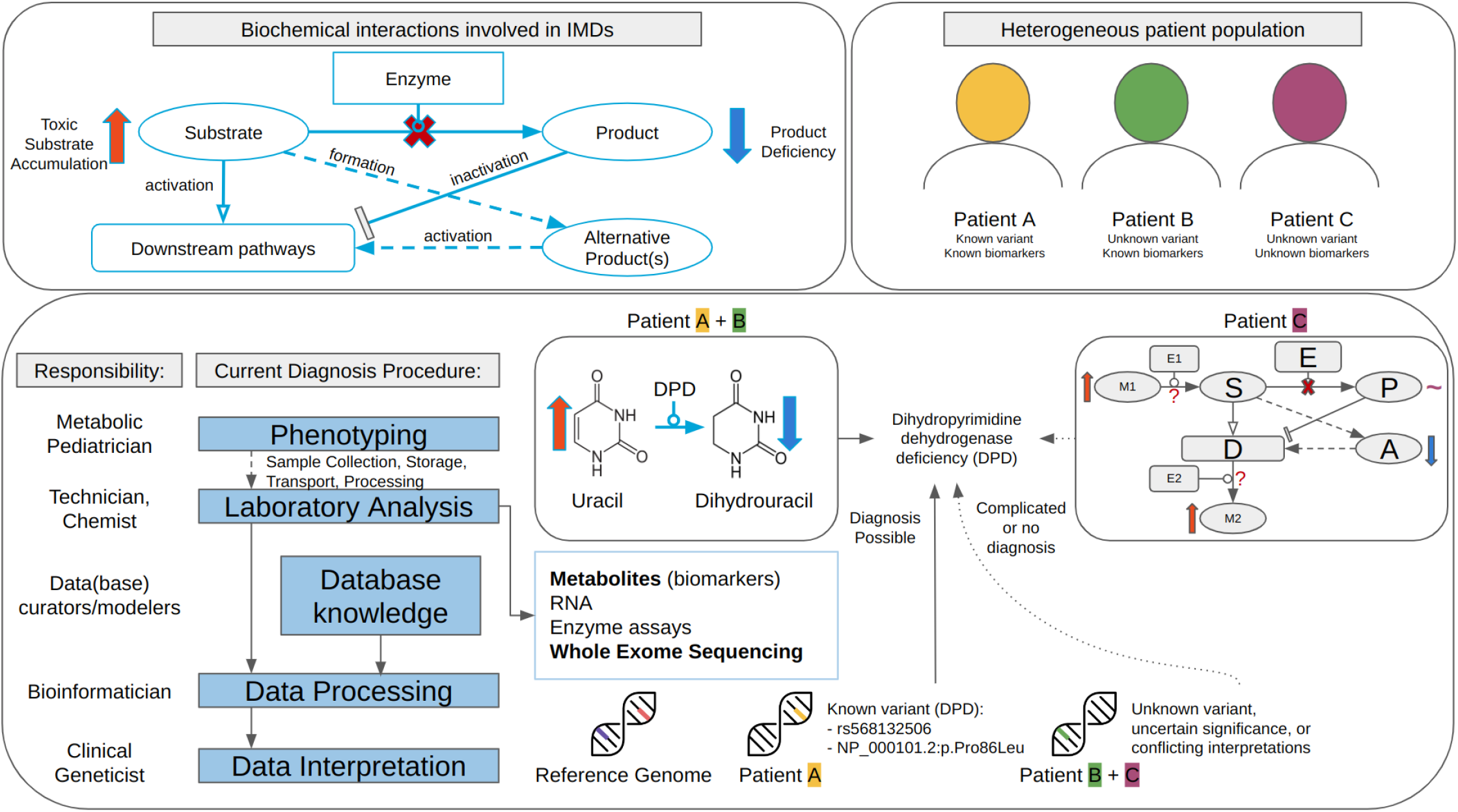
Overview of biochemical interactions involved in IMDs (top left); the current diagnostic procedure, and challenges in diagnosis using targeted metabolite or WES data for heterogeneous patient populations.

Unfortunately, diagnosing IMDs using metabolites can be challenging due to the commonly observed overlap between biomarkers, since the individual compounds are often involved in more than one metabolic pathway and can therefore be metabolized to various products. Furthermore, the diagnostic process can be quite time-consuming, requiring a manual inspection by an expert in the field, who needs to be familiar with all relevant metabolic conversion and their respective enzymes to point out the malfunctioning protein. Last, current clinical diagnoses are lacking a visualization of the connections between individual metabolic biomarkers and the enzymes involved in their synthesis and degradation.

Therefore, this study provides a proof-of-concept framework for the integration of metabolic interactions knowledge with clinical patient data, and identifies current challenges for scaling up this approach. We hypothesize that the combination of this knowledge and patient data in one visualization can aid in the diagnosis of IMDs, by providing an overview of the processes relevant to the patient-specific deficient protein. With this approach, the attention progresses from individual markers to changes at the process level, which enables linking biological pathway knowledge to clinical cases. This direct link shows which metabolic reactions are disturbed, which proteins are related to these reactions, and potentially which specific protein is impaired, aiding diagnosis. Furthermore, metabolic disturbances can be recognized which cannot be attributed directly to the disorder, revealing potential blind spots in existing clinical knowledge.

Our framework was tested on two groups of IMDs, pyrimidine metabolism and the urea cycle, with a well-understood molecular mechanism known for biomarker overlap for several IMDs due to their common metabolite carbamoyl phosphate [9]. Furthermore, pyrimidine disorders often present with nonspecific clinical symptoms and a lack of a clear genotype-phenotype correlation [10–12], while urea cycle disorders are often more specific (e.g. hyperammonemia, lethargy, vomiting, coma) [13].

The presented framework highlights chances for the IMD field as a whole regarding data integration and reuse, by showcasing that improving data and identifier (ID) harmonization increases the integration of clinical data with pathway knowledge and biomarker information. Furthermore, the framework could aid in the diagnostic process of other (novel) IMDs and is adaptable to analyze different types of IMDs and functional assays in the future, as well as integrating other types of (omics) data analysis, e.g. transcriptomics, metabolomics, and fluxomics. By using visualization techniques from common network approaches, the framework could also be extended with information on drug targets or genetic variants, which could allow for personalized medicine. Last, since this study combines several research fields and demonstrates an interdisciplinary approach, this paper will address each field individually with the hope of closing the gap between the data collection and interpretation, data curation and modeling, and data processing and interoperability.

## 1. Methods

### Workflow

Figure 2 shows the proposed workflow to connect clinical data to pathway models and theoretical biomarker data. Knowledge from various databases had to be integrated into the framework, which is summarized in Table 1. All data processing steps were captured in an RMarkdown script [14] in the R programming language (version 4.1.3) [15], tested through RStudio (version 2022.02.2) [16], available at https://github.com/BiGCAT-UM/IMD-PUPY.

**Table 1:** Overview of Databases and tools used within the workflow

| Database/tool [reference] | Description | Workflow Step |
| --- | --- | --- |
| ChEBI [19] | Chemical Entities of Biological Interest (ChEBI) is a freely available dictionary of molecular entities focused on 'small' chemical compounds. | 1-3, 5-7 |
| Wikidata [20] | Wikidata is a free and open knowledge base that can be read and edited by both humans and machines | 1 |
| UHPLC-MS/MS [18] | Ultra High-Performance Liquid Chromatography-Tandem Mass Spectrometer is a chemical technique combining the physical separation capabilities of liquid chromatography with the mass analysis capabilities of mass spectrometry. | 1 |
| PathVisio [21] | PathVisio is a free open-source pathway analysis and drawing software that allows drawing, editing, and analyzing biological pathways. | 2 |
| UniProt [22] | UniProt is a freely accessible database of protein sequence and functional information, many entries being derived from genome sequencing projects. | 2 |
| Rhea [23] | Rhea is an expert-curated knowledge base of chemical and transport reactions of biological interest - and the standard for enzyme and transporter annotation in UniProtKB. | 2 |
| OMIM [24] | Online Mendelian Inheritance in Man (OMIM) is a comprehensive, authoritative compendium of human genes and genetic phenotypes that is freely available. | 2 |
| WikiPathways [25] | WikiPathways is a database of biological pathways maintained by and for the scientific community. | 2, 4, 6, 7 |
| WikiPathways SPARQL Endpoint [27] | The semantic web format of the WikiPathways database, using the Resource Description Framework (RDF) as data format which can be queried through the SPARQL-query language. | 2, 4, 6, 7 |
| HGNC (gene ID) [60] | The HGNC is a resource for approved human gene nomenclature. | 4 |
| HMDB [30] | The Human Metabolome Database (HMDB) is a freely available electronic database containing detailed information about small molecule metabolites found in the human body. | 4, 5 |
| Cytoscape (REST API) [32] | Cytoscape is an open-source software platform for visualizing complex networks and integrating these with any type of attribute data. cyREST is a language-agnostic, programmer-friendly RESTful API module for Cytoscape, allowing for programmatic access to Cytoscape features. | 7 |
| GitHub (github.com) | GitHub is a web-based hosting service for version control using git. | 1-8 |

**Figure 2:**
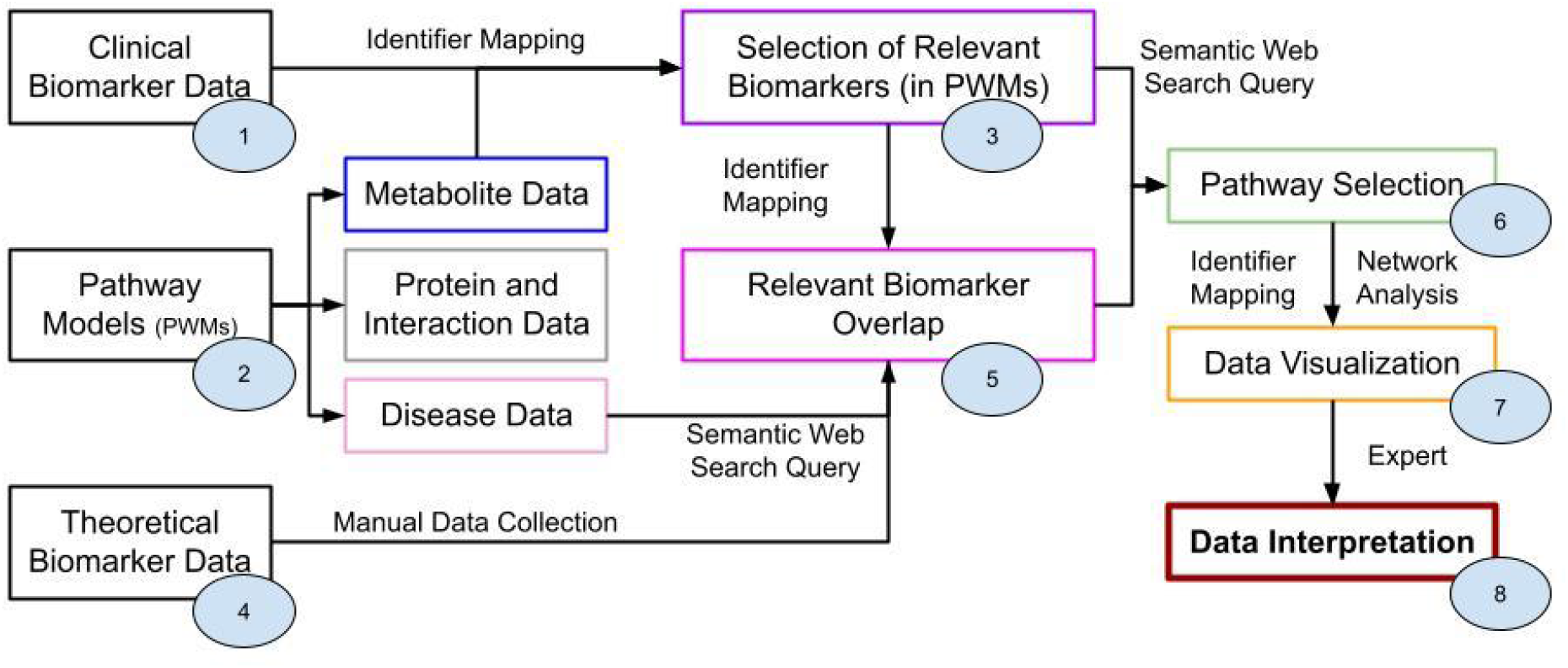
Depiction of workflow to interpret clinical data of IMDs through network analysis; each circle refers to a specific step within the Materials and Methods Section.

### Clinical Biomarker Data

Biomarker data from 22 patients previously diagnosed with a pyrimidine or urea cycle IMD was collected through two targeted chemical assays in urine [17,18]; metabolite concentrations were reported in μmol/mmol creatinine and patient age in months. Four patients were removed from this study, due to missing data for the AA panel (patients labeled B, C, P, and Q). The same assays were used to collect reference data from other patients suspected of having an IMD, however with no apparent IMD as assessed by selective metabolic screening. Reference data for purines and pyrimidines (PUPY panel) included 4853 samples selected over ten years; amino acids (AA panel) 1872 samples over five years. The reference data was categorized in five age categories; data from the overarching category 0 to 16 years was used if no reference value was available for a specific age category. For the 88 chemical biomarkers present in the patient data, four were disregarded from further data analysis due to missing reference data: n-carbamyl-aspartate (CHEBI:32814), allantoin (CHEBI:15676), cytosine (CHEBI:16040), and cytidine (CHEBI:17562). The patient and reference data was annotated with corresponding ChEBI [19] identifiers (IDs) or Wikidata [20] IDs when no ChEBI ID was available. Patient data was anonymized and five biomarkers were disregarded: allopurinol (used as treatment) and its metabolite oxypurinol; argininosuccinic acid anhydride (ASA-anhydride) (obsolete after switching the separation method from anion exchange chromatography to UHPLC-MS/MS for AA analysis [18]); and CysHCys and 2,8-dihydroxyadenine (metabolites without a ChEBI ID). Table 2 details the sample size, diseases, and corresponding age ranges used in this study. No patients or their caregivers have objected to the anonymous use of their leftover material from routine diagnostics for laboratory development and validation purposes.

**Table 2:** Sample size sorted on disease classification and age range. The patient labels (A to T) are included per relevant age category. The HGNC name for the gene linked to the disorder is provided if deviating from the common abbreviation. The ^#^ indicates if a patient has received treatment.

| Age range (years) →<br>Diseases ↓ | 0-1 | 1-5 | 5-16 | 16+ |
| --- | --- | --- | --- | --- |
| *Pyrimidine* - Full name (Abbreviation, OMIM-ID, HGNC name) |  |  |  |  |
| Beta-ureidopropionase deficiency (UPB1D, 613161, UPB1) | 1 (J) |  |  |  |
| Dihydropyrimidine dehydrogenase deficiency (DPD, 274270, DPYD) | 1 (I) | 1 (A) | 1 (E) | 1 (K) |
| *Urea cycle* - Full name (Abbreviation, OMIM-ID, HGNC name) |  |  |  |  |
| Argininosuccinic aciduria (ASLD, 207900, ASL) |  | 2 (M, N) | 2 (L, T) | 1 (F) |
| Citrullinemia Type I (ASS1, 215700) | 1 (D) |  |  |  |
| Hyperornithinemia-hyperammonemia-ho mocitrullinuria syndrome (HHHS, 238970, SLC25A15) |  |  | 1 (H#) |  |
| Ornithine Transcarbamylase deficiency (OTCD, 311250, OTC) |  | 1 (G) | 2 (R, S) | 1 (O) |

### Pathway models

Since the clinical patient data also included metabolites from the purine pathway, IMDs in this group were added to the analysis to serve as control data points. Machine-readable versions of the purine, pyrimidine, and urea cycle metabolic pathways were created using the pathway editor and curation tool PathVisio (version 3.3.0) [21], as well as pathway models (PWMs) on biomarkers, visualizing several markers missing from the main pathway models. All proteins were annotated with UniProt IDs [22], and directed Rhea IDs [23] for the metabolic conversions. Corresponding ChEBI IDs [19] from Rhea were used to annotate the substrate and product metabolites. IMDs were annotated with OMIM disease IDs [24]. Data on the created PWMs was deposited in WikiPathways [25] and retrieved from RDF data format (Resource Description Framework [26]) through the WikiPathways SPARQL endpoint [27] (data from September 2021 [28]).

### Selection of Relevant Biomarkers (in PWMs)

All biomarkers were compared to the lower or upper reference values; below the lower limit indicated a decrease (negative change) and above the upper limit indicated an increase (positive change). Biomarker values in between or exactly equal to the reference values were designated as unchanged. Missing biomarker data (null-values) were disregarded, as well as patient or reference concentration data being equal to zero. All resulting calculated values were log(2) transformed to show proportional changes, resulting in a log2FC. The changed biomarkers were compared against existing PWMs to find missing entries through the WikiPathways SPARQL endpoint (data from September 2021).

### Theoretical Biomarker Data

Since the chemical assay for pyrimidine metabolites also measures purine compounds, we collected theoretical biomarker data for both pathways, in addition to the urea cycle. Potentially relevant biomarkers for these disorders were retrieved manually from IEMbase [29] V 2.0.0 (accessed on 2021-08-05) through their HGNC gene name as HMDB IDs [30], including the sample matrix, and positive or negative concentration change. The latter was converted to a numeric scale for each of the five provided age categories. The biomarkers in IEMBase were represented through arrows (and some other characters) to show relative increases or decreases rather than numeric values. These visualizations were converted to a numeric scale (from -3 to +3) according to these rules:

| Visualization | Conversion | Visualization | Conversion | Visualization | Conversion |
| --- | --- | --- | --- | --- | --- |
| ↑↑↑, ↑↑, ↑ | +3, +2, +1 | ↑ - ↑↑, ↑↑ - ↑↑↑ | +1.5, +2.5 | n to 1, n to 2 | +0.5, +1.5 |
| ↓↓↓, ↓↓, ↓ | - 3, -2, -1 | ↓-↓↓, ↓↓-↓↓↓ | -1.5, -2.5 | n, +/- | 0 |

Correlations between individual metabolic biomarkers and diseases were visualized in a heatmap (Euclidean distance) with the gplots package (version 3.1.1, https://cran.r-project.org/package=gplots); positively changed biomarkers were colored red (using three shades to show mildly, high, and very high), negatively changed markers blue (again in three shades); markers which were not altered for a disease were colored white. Disorders without any biomarker data for a specific age category were removed from the visualizations.

### Relevant Biomarker overlap

All biomarkers were manually linked from ChEBI IDs (patient and pathway model data) to their corresponding HMDB IDs (theoretical biomarker data). The patient biomarker data was converted to the same scale as the theoretical biomarkers (values for log2FC above 3 or below -3 were set at 3 and -3, respectively). The patient data was visualized together with the theoretical biomarker visualization, removing small changes (log2FC between -0.05 and 0.05).

### Pathway Selection

Relevant pathways were found through a query against the WikiPathways SPARQL endpoint matching the changed biomarkers. The pathways were sorted based on the highest number of matching biomarkers. A maximum of three pathways were selected, based on including most unique biomarkers.

### Data visualization

The data for each patient was visualized with the network analysis tool Cytoscape [31] (version 3.9.1), by using the Cytoscape REST API [32] (version v1) and WikiPathways App for Cytoscape [33] (version 3.3.10) through R. The absolute highest value for the log2FC was used to determine the color scale, using a five-point scale to accommodate for small changes (values between -1.5 and 1.5) and high (abnormal) biomarker values. If no value was available for a node within the network, the fill color was set to gray.

### Data Interpretation

For each patient, the network data visualization (framework step 7) and relevant biomarker overlap heatmap (step 5) were provided to two Laboratory Specialists in Biochemical Genetics, after which narrative feedback on a potential diagnosis was collected.

## 2. Results

A framework was designed to visualize clinical biomarker data for IMDs through their metabolic interactions. In order to explain the findings of this interdisciplinary study, this section is divided in three paragraphs, so that experts from different research fields can directly find the information most relevant to them, while also being able to switch outside of their expertise.

### Clinical geneticists, metabolic pediatricians, biologists, and chemists

This group of experts is mainly responsible for the data collection and interpretation (e.g. Metabolic Pediatricians, Laboratory Specialist), and are involved at the direct start of the diagnostic pipeline and the final diagnostic step. Our framework was tested on data from 16 patients with a variety of pyrimidine and urea cycle IMDs and is summarized in Table 2. In total 88 clinical markers were measured in urine samples, 34 through the PUPY panel (purines and pyrimidines) and 54 by the AA panel (amino acids). Theoretical biomarkers for the investigated phenotypes were obtained from an online database (IEMbase), finding 27 unique metabolic biomarkers relevant for urine samples. Table 3 shows the number of (significantly altered) biomarkers linked to reference data for each patient, as well as the number of biomarkers found in a metabolic pathway. Two laboratory specialists in biochemical genetics used the data visualizations from our framework to arrive at an IMD diagnosis, by combining the heatmap showing theoretical biomarkers and related enzymes with the network biomarker data visualization.

**Table 3:**
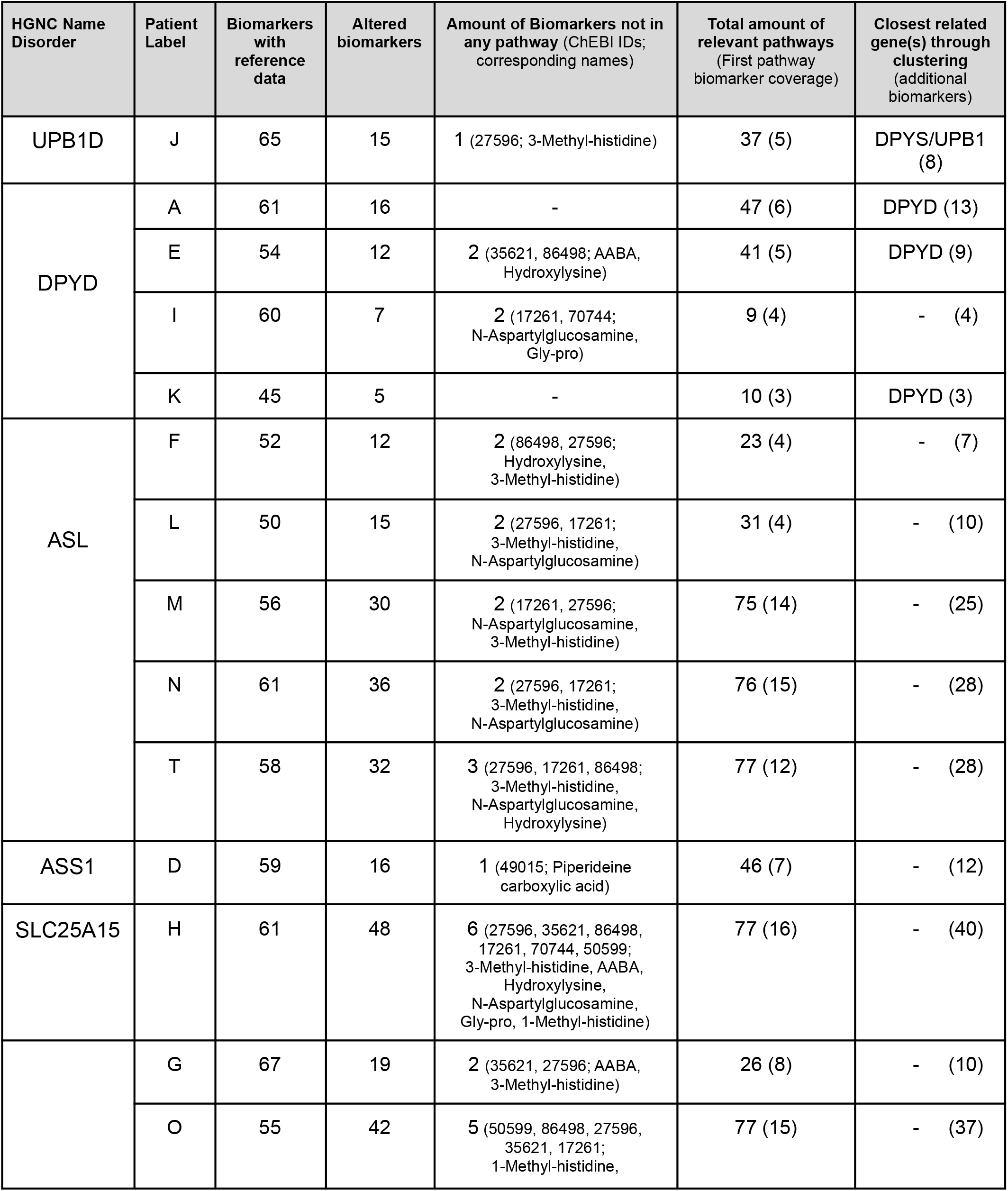
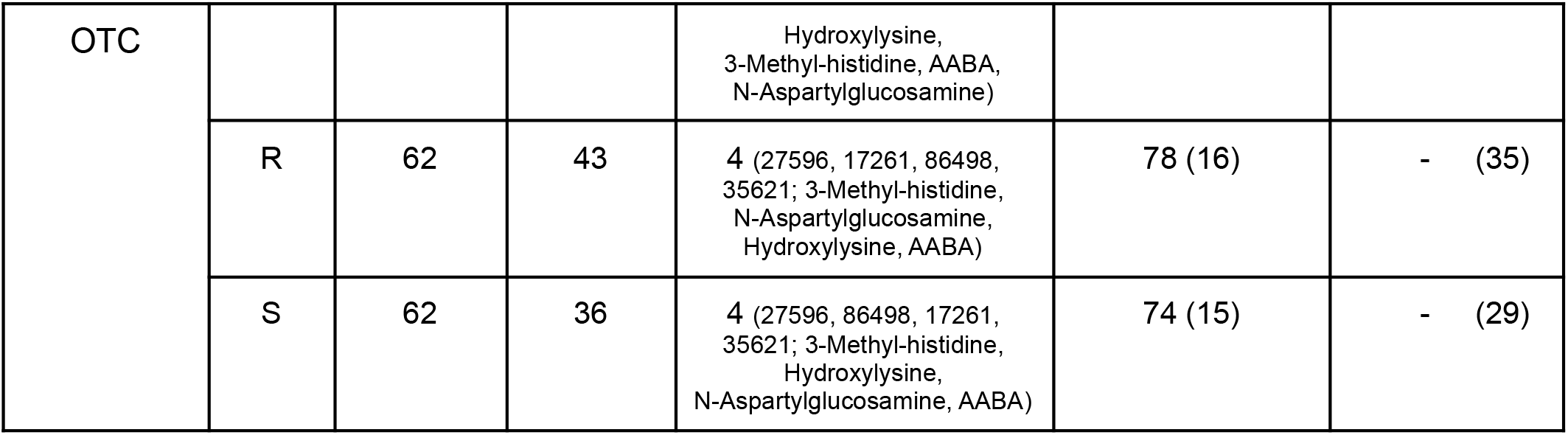
Overview of biomarker data per patient, including number of altered biomarkers and coverage thereof in pathways models, as well as theoretical biomarker clustering information for the investigated phenotypes and corresponding genes (‘-’ indicates that no closely related gene could be found through clustering).

Figure 3 shows the theoretical biomarkers for their respective IMD class (purine, pyrimidine, urea cycle) and data for one patient (age category 0-1 year, diagnosed originally with DPYD, in purple, labeled patient I) as a heatmap. Comparing theoretically changed biomarkers to patient data is the first step in selecting potentially relevant phenotypes and affected proteins, and can be used to imply which biochemical reactions or pathways are disturbed. Rows indicate individual phenotypes (right axis), and are clustered (left axis) based on their overlapping biomarker profiles (bottom axis). The top left of Figure 3 shows that for example the first two rows representing SLC25A15 and OTC (both urea cycle disorders) are clustered together, due to their overlapping biomarkers orotic acid (HMDB0000226) and homocitrulline (HMDB0000679). However, for SLC25A15 an excessive amount of homocitrulline is produced and a small increase in orotic acid can be noted, while for OTC both metabolites are increased in a similar amount. Disorders clustered together can be difficult to diagnose, due to marginal changes in or low numbers of known biomarkers, and overlap between the markers. The sample obtained from patient I showed four additionally changed biomarkers compared to the theoretical values; however no direct relation to the theoretical biomarker profile of DPYD was observed.

**Figure 3:**
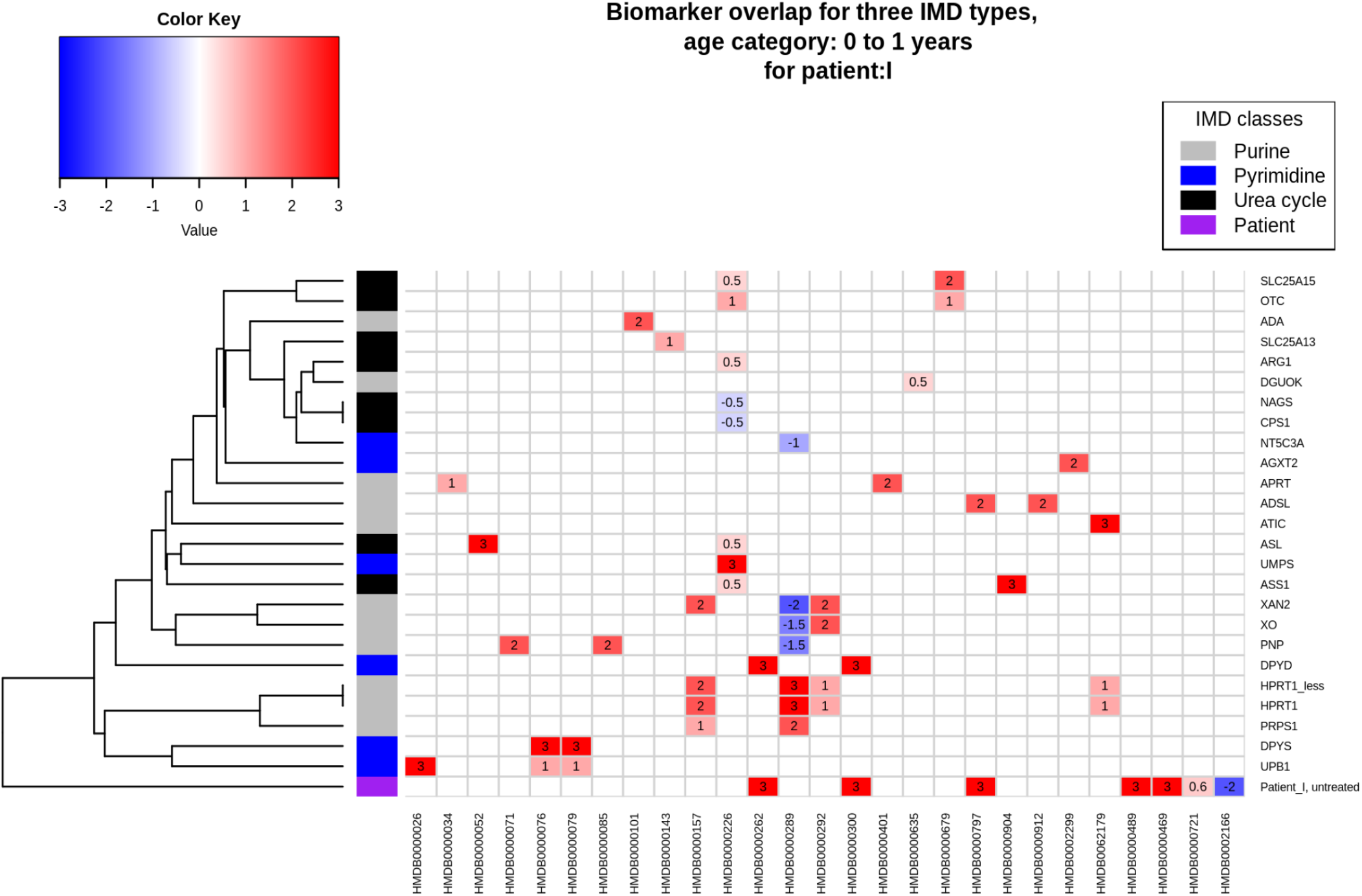
Visualization of theoretical overlap between biochemical urine markers for individual IMDs in the purine, pyrimidine, and urea cycle pathways. Rows are linked to individual phenotypes (right axis), and clustered (left axis) based on their overlapping biomarker profiles (bottom axis). Protein names correspond to the following disorders: ADA: Adenosine deaminase deficiency, SLC25A13: Citrin deficiency, ARG1: Arginase deficiency, DGUOK: Deoxyguanosine kinase deficiency, CPS1: Carbamoyl phosphate synthetase I deficiency, NAGS: N-Acetylglutamate synthase deficiency, NT5C3A: Pyrimidine 5-nucleotidase superactivity, AGXT2: Beta-aminoisobutyrate-pyruvate transaminase deficiency, SLC25A15: Ornithine transporter deficiency, OTC: Ornithine transcarbamylase deficiency, APRT: Adenine phosphoribosyltransferase deficiency, ADSL: Adenyl-succinate lyase deficiency, ATIC: AICAr transformylase/IMP cyclohydrolase deficiency, UMPS: Orotic aciduria type I, ASL: Argininosuccinic aciduria, ASS1: Citrullinemia type I, XAN2: Xanthinuria, Type II, XO: Xanthinuria, Type I, PNP: Purine nucleoside phosphorylase deficiency, HPRT1_less: Kelley-Seegmiller syndrome, HPRT1: Lesch-Nyhan syndrome, PRPS1: Phosphoribosyl pyrophosphate synthetase 1 superactivity, DPYD: Dihydropyrimidine dehydrogenase deficiency, DPYS: Dihydropyrimidinase deficiency, UPB1: Beta-ureidopropionase deficiency HMDB IDs resemble these metabolites: HMDB0000026: N-Carbamyl-beta-alanine, HMDB0000034: Adenine, HMDB0000052: Argininosuccinate, HMDB0000071: Deoxyinosine, dIno, HMDB0000076: Dihydrouracil, HMDB0000079: Dihydrothymine, HMDB0000085: Purine nucleoside phosphorylase, HMDB0000101: Deoxyadenosine, HMDB0000143: Galactose, HMDB0000157: Hypoxanthine, HMDB0000226: Orotic acid, HMDB0000262: Thymine, HMDB0000289: Uric acid, HMDB0000292: Xanthine, HMDB0000300: Uracil, HMDB0000401: 2,8-Dihydroxyadenine, HMDB0000635: Succinylacetone, HMDB0000679: Homocitrulline, HMDB0000797: SAICA riboside, HMDB0000904: Citrulline, HMDB0000912: Succinyladenosine, HMDB0002299: beta-aminoisobutyrate, HMDB0062179: AICA riboside. Additional biomarkers relevant for this specific patient (not part of IEMbase data): HMDB0000489: N-Aspartylglucosamine, HMDB0000469: 5-(Hydroxymethyl)uracil, HMDB0000721: Gly-pro, HMDB0002166: (S)-Beta-aminoisobutyrate.

Regarding all patients, the biomarker profile of only four patients clustered with a potential gene of interest (Table 3 last column), with three patients (labeled A, E, K) closely resembling the theoretical biomarkers for their corresponding disorder DPYD. Interpreting the remaining patient data with knowledge of the biochemical interactions between the biomarkers is needed to arrive at a diagnosis.

Figure 4 shows the data visualization for patient I on the pathways selected for this patient: ‘Biomarkers for pyrimidine metabolism disorders’ (left) and ‘Purine metabolism’ (right) pathways. As expected for DPYD, the pyrimidine pathway which includes the DPD protein shows the most relevant metabolic changes for this patient. Two metabolites (thymine, uracil) which are directly converted by DPD show elevated levels; one direct downstream metabolite of thymine (5-OH-methyluracil) also shows a high concentration. Two downstream metabolites of DPD (dihydrouracil, beta-alanine) are found to be within the healthy reference values, whereas (S)-beta-aminoisobutyrate also downstream of DPD shows a decreased concentration. The second selected pathway on purine metabolism shows elevated levels of SAICA-riboside, which was not expected for this disorder and might suggest physiological immaturity.

**Figure 4:**
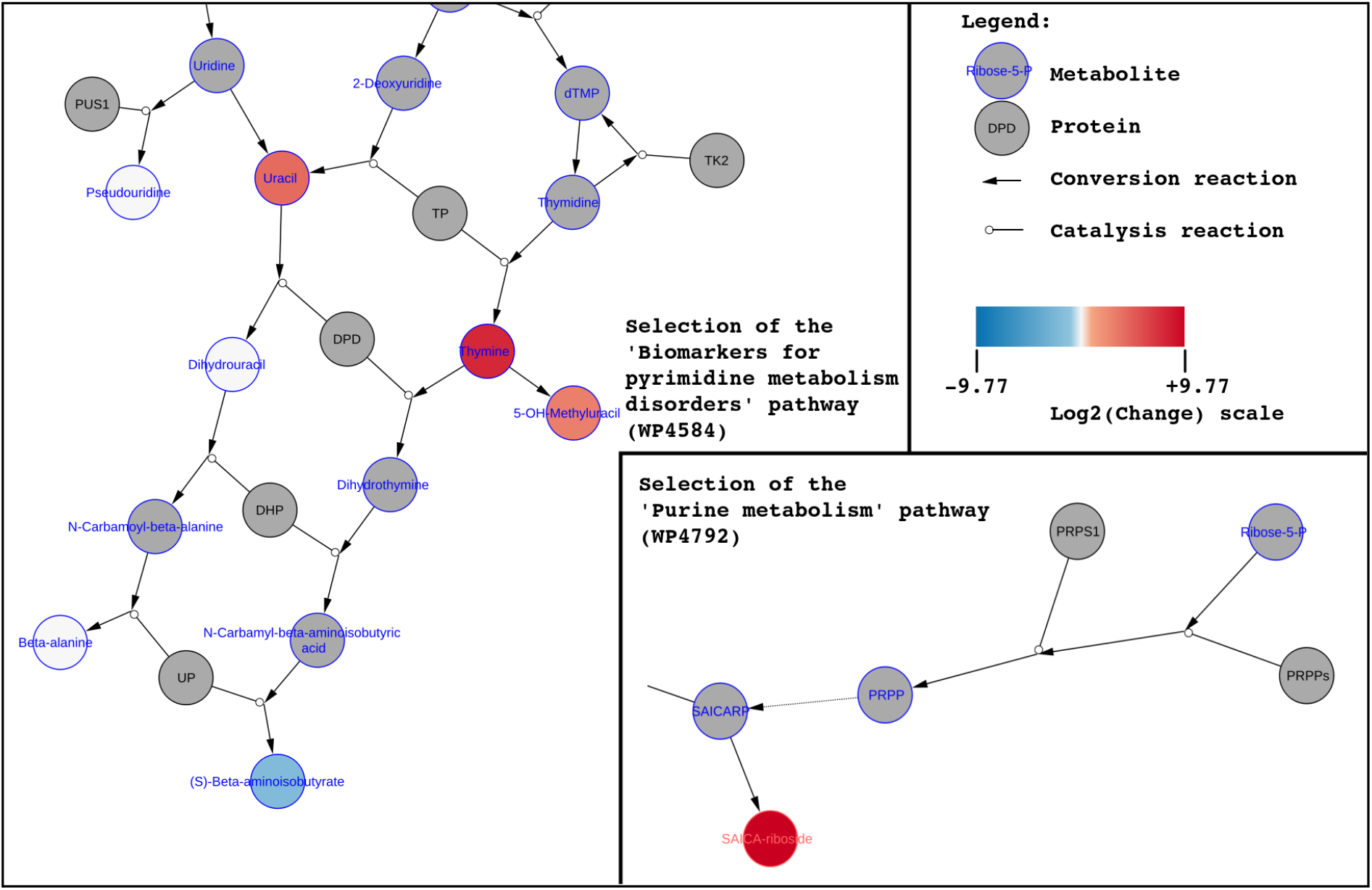
Network visualization of biochemical markers for patient I (diagnosed with DPYD). *Left:* selected section of pyrimidine biomarkers pathway; *Right:* selection of purine metabolism pathway.

In total, nine disorders out of the 16 patient samples were diagnosed with the correct IMD, whereas for four patients the visualization suggested further informative assays (see Table 6). These latter patients included one case of Dihydropyrimidine dehydrogenase deficiency (DPYD - patient E), and three cases of Ornithine Transcarbamylase deficiency (OTC - patients G, R, and S). Samples from patients under treatment, e.g. patient H (diagnosed with hyperornithinemia-hyperammonemia-homocitrullinuria (HHH) syndrome, also known as ornithine translocase (SLC25A15) deficiency) receiving citrulline, were difficult to diagnose since the framework cannot distinguish between abnormal biomarker values due to treatment or caused by the IMD. Patient J (diagnosed with Beta-ureidopropionase deficiency, UPB1D) was not correctly diagnosed by both experts, which we attribute to the very mild disturbances in the biomarker patterns. Last, patient O (diagnosed with Ornithine Transcarbamylase deficiency, OTC), showed an unexpectedly higher value for arginine rather than ornithine.

### Data(base) curators and modelers

This section describes the data curation and modeling aspects of this study; often an invisible layer in the diagnostic process however an important influence on the results of a diagnosis. Out of the 88 biomarkers measured through the targeted metabolic assays, two could not be annotated with one unified database ID from ChEBI. Six new pathway models were created for this project, to provide interoperability between the metabolic interactions relevant for the studied IMDs and the clinical biomarker data. Table 3 details how many relevant clinical biomarkers were missing from any PWM from the WikiPathways database (framework step 3), the pathway’s coverage of biomarkers relevant for each patient (pathway), and the highest total of markers covered by one pathway. Table 4 describes the content of each pathway, regarding proteins, metabolites, interactions, and described disorders. There were seven markers not part of any pathway model (previously existing or newly created) with the corresponding ChEBI IDs: 28315, 40279, 17755, 43433, 89698, 49015, 61511, which can be used for future curation. The available theoretical reference data for urine samples with a database identifier (HMDB) left 23 unique biomarkers, linked to 25 individual IMD phenotypes. Nine disorders were missing theoretical urinary biomarker data, and for one disorder the molecular mechanism is still unclear, therefore missing a specific protein connection.

**Table 4:** Amount of unique DataNodes (proteins, metabolites) and disorders captured by each pathway model. Uniqueness counts based on unification to HGNC names (Proteins), ChEBI IDs (Metabolites), and OMIM URLs (Disorders).

| Model Name | PWM ID | #Proteins<br>(HGNC) | #Metabolites<br>(ChEBI) | #Interactions<br>(Rhea) | #Disorders<br>(OMIM) |
| --- | --- | --- | --- | --- | --- |
| Purine metabolism and related disorders | WP4224 | 22 | 61 | 51 | 17 |
| Pyrimidine metabolism and related diseases | WP4225 | 17 | 41 | 33 | 15 |
| Urea cycle and related diseases | WP4571 | 9 | 16 | 7 | 9 |
| Urea cycle and associated pathways | WP4595 | 24 | 35 | 23 | 12 |
| Biomarkers for urea cycle disorders | WP4583 | 12 | 29 | 10 | - |
| Biomarkers for pyrimidine metabolism disorders | WP4584 | 15 | 39 | 29 | - |

### Programmers, data scientists, and bioinformaticians

The last group addressed in this section provides the glue that holds the analysis section of the diagnostic pipeline together, and is responsible for data processing and interoperability. Theoretical biomarker data was collected for all IMDs in the pyrimidine and urea cycle pathway; biomarkers for the purine pathway were included to represent true negative values. In total, 17 purine, 10 pyrimidine, and 8 urea cycle IMDs were included (35 disorders in total) in the PWMs. The annotated data from framework step 1 (Figure 2) was used to find relevant pathways for visualization, which led to 171 pathways in total, including one or more distinct biomarkers. Fifteen of these pathways contain 10 or more markers, displayed in Table 5, which could potentially be ideal candidates for the data visualization.

**Table 5:** A total of fifteen pathways contain a high amount of biomarkers for both assays, with at best 46 markers included in one individual pathway model. ^%^-sign indicates which pathways were not found in the top three relevant pathways for any patient.

| Pathway ID | Title of Pathway | ChEBI IDs In PW |
| --- | --- | --- |
| WP3604 | Biochemical pathways: part I% | 46 |
| WP4042 | Nucleobase catabolism% | 21 |
| WP4225 | Pyrimidine metabolism and related diseases | 18 |
| WP1936 | Transport of inorganic cations/anions and amino acids/oligopeptides% | 17 |
| WP4584 | Biomarkers for pyrimidine metabolism disorders | 17 |
| WP1935 | Transport of bile salts and organic acids, metal ions and amine compounds% | 17 |
| WP3925 | Amino acid metabolism | 15 |
| WP4224 | Purine metabolism and related disorders | 14 |
| WP4082 | Nucleotide salvage<sup>%</sup> | 14 |
| WP1938 | tRNA aminoacylation<sup>%</sup> | 13 |
| WP1937 | Transport of vitamins, nucleosides, and related molecules<sup>%</sup> | 12 |
| WP4792 | Purine metabolism | 12 |
| WP4595 | Urea cycle and associated pathways | 11 |
| WP4022 | Pyrimidine metabolism | 10 |
| WP4583 | Biomarkers for urea cycle disorders | 10 |

By only querying the pathway data for relevant biomarkers (instead of all markers in a panel), a customized visualization was created for each patient. The selection of the top three pathways containing the highest number of unique markers was performed using SPARQL-queries. One pathway for three patients was selected manually, aiming to include relevant metabolic interaction containing biomarkers with the largest change. Table 6 shows which pathways ended up in the top three for each patient, as well as which biomarkers were not part of the visualization after selecting the top three.

**Table 6:**
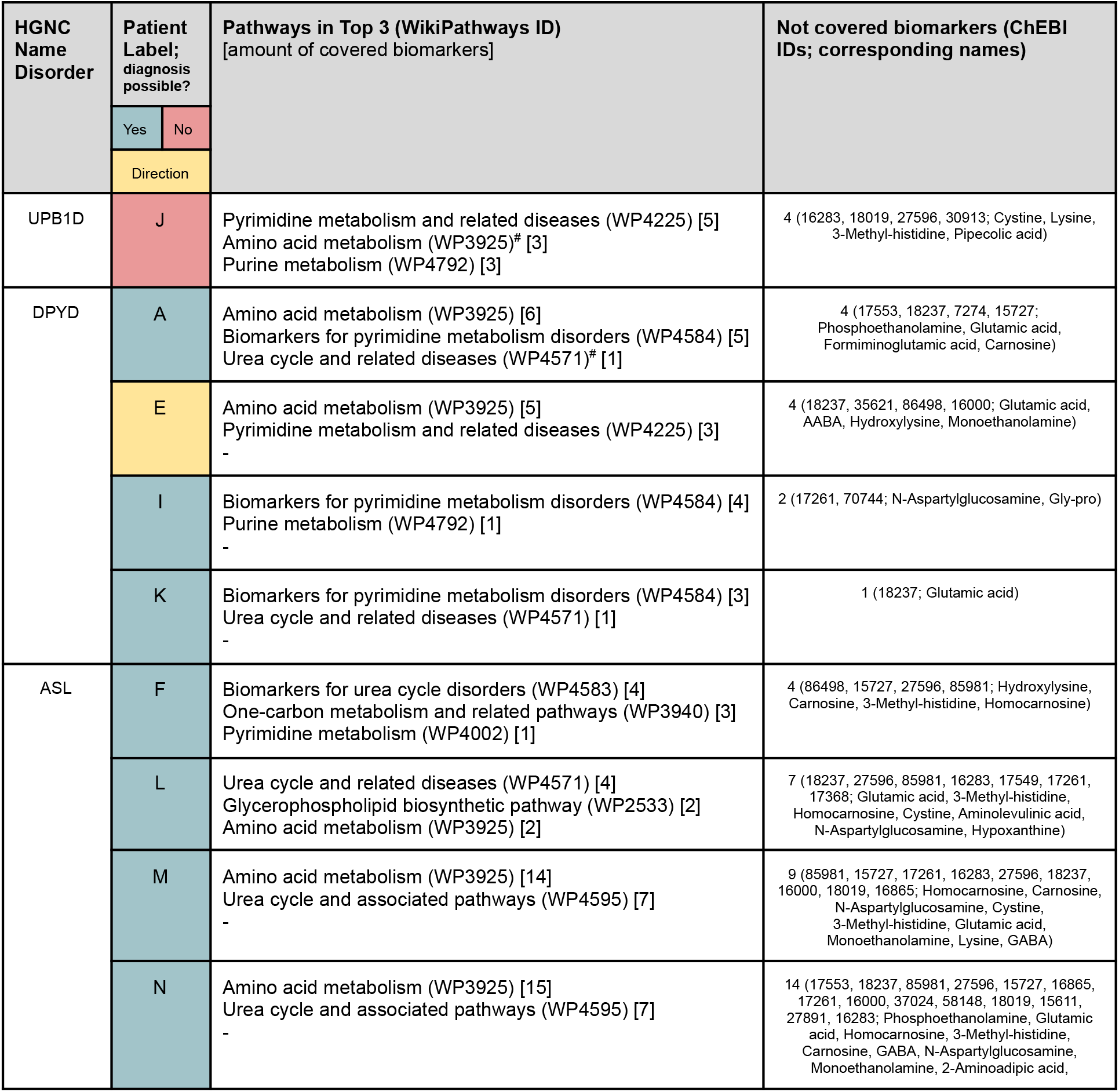
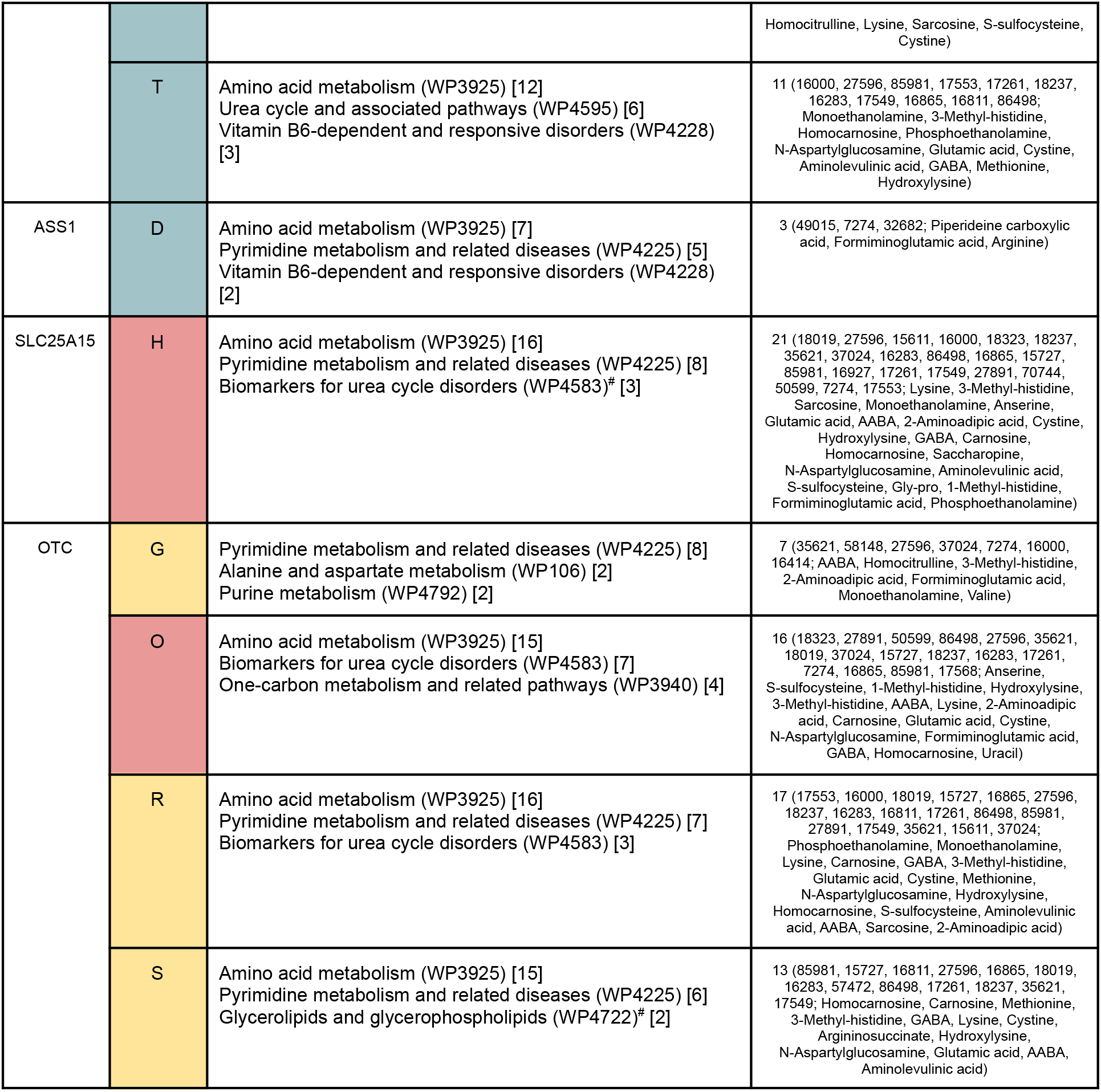
Top three pathways containing most unique biomarkers, as well as which biomarkers were not covered by the top three pathways (however were part of a PWM). ^#^ indicates different pathways cover the same amount of unique metabolites leading to a manual selection of the most relevant pathway. - indicates no third pathway was selected.

## 3. Discussion

Our proposed framework is based on the combination of clinical data, online available biomarker information, and metabolic reaction models. This framework creates the possibility to visualize clinical biochemical data on the pathway level, allowing for a more detailed interpretation of the connections between the different markers. The developed framework was designed for individual patient analysis and optimized for pyrimidine and urea cycle disorders with biomarkers measured through targeted assays. We believe this framework can be extended to other IMDs and more biological matrices. Also, several challenges should be taken into account before scaling up the framework. This section is again divided in three paragraphs, to describe the challenges for data collection and interpretation, data curation and modeling, and data processing and interoperability.

### Clinical geneticists, metabolic pediatricians, biologists, and chemists

Our developed framework enables the visualization of clinical biomarker profiles with biological pathway knowledge, by connecting individual markers to changes on the process level. This approach shows which metabolic reactions are disturbed, which proteins are related to these reactions, and potentially which specific protein is impaired, aiding diagnosis. Existing data interpretation approaches often require a manual inspection of pathways and interactions, which do not include the clinical data. Furthermore, metabolic disturbances can be recognized which cannot be attributed directly to the disorder, revealing potential blind spots in existing clinical knowledge. However, the data integration needed for this approach requires database identifiers; therefore we advise the (rare) disease community to include these identifiers (from a publically available database) for each compound measured through a metabolic assay. The developed framework is extendable with in-house biomarker data, knowledge from other databases or literature, and additional data from blood samples or other relevant matrices. Even though recent advances in clinical urinary biomarker measurements [17] have aided in the diagnosis of some IMDs, most markers are currently not used for newborn screening [34] due to limited detectability of these biomarkers in dried blood. The inclusion of an additional matrix could provide a broader overview of the metabolic disturbances in a patient and lead to a more comprehensive isolation of the involved metabolic interactions. The framework also leaves room for manual selection of potentially relevant pathways by experts, which could be aided by reviewing the patient-specific heatmap which visualizes theoretical biomarkers. Our framework could be enhanced by selecting the top three pathways covering the most unique biomarkers while prioritizing the markers with the highest log2FC. Currently, diagnostic laboratories for inborn errors of metabolism often report exact metabolite concentrations in diagnostic patient reports; Z-scores are (rarely) used as a measure to compare patient values to a control population. The pathway and network models used in our framework normally report (log)-fold changes (log2FC) to compare two groups (diseased versus control) or z-scores as a measure for over-representation of pathway entities such as metabolites. We believe that laboratory specialists would benefit from learning to interpret this type of data and visualization as a new diagnostic tool. In this study, the network model helped to easily diagnose 9 out of 16 patient samples, and pointed in the correct direction or suggested follow-up analysis for 4 patients. These numbers are similar to the original diagnostic outcome for the metabolite pipeline; the remaining 7 out of 16 patients could previously only be diagnosed with additional tests (e.g. protein loading test, WES, clinical information). Patient H (previously diagnosed with ornithine translocase (SLC25A15) deficiency with the main biomarker homocitrulline, HMDB0000679) was found difficult to recognise, both through the original metabolic pipeline and our framework. This issue was most likely due to the administered treatment with citrulline, highlighting the importance of not only clinical but also medication information for a proper diagnostic workflow. As in any other computational framework, only data from untreated persons should be used. For the validation of a recently implemented diagnostic tool called targeted urine metabolomics (TUM) [35], similar samples were used as discussed in our study. When comparing the data interpretation from TUM and the developed framework, we can deduce that comparable interpretations were reached. This agreement was also found for the Ornithine Transcarbamylase deficiency (OTC) cases, which remains difficult to diagnose in women since the disorder is X-linked causing an atypical biomarker pattern. Patient O is an example of such, where we hypothesize that the cyclic metabolic conversion of arginine, argininosuccinate, and ornithine into one another could be the cause of this unexpected pattern. To understand these atypical cases of OTC and corresponding biomarker patterns, data from more patients is required and we recommend other laboratories to share their data on IMDs if possible. Sharing (more) rare disease patient data would also help to understand the effects of ethnicity, age range, or sex on the molecular mechanism of IMDs. For future studies, the interest should shift to measuring metabolite fluxes [2] over a longer timespan to better understand how for example protein intake triggers decompensation [36].

### Data(base) curators and modelers

The presented framework highlights chances for the IMD field as a whole regarding data integration and reuse, one of the cornerstones of data modeling. Our framework leverages on data and identifier (ID) harmonization to increase the machine readability of existing IMD data. This harmonization was required for the integration of clinical data with pathway knowledge and biomarker information. All metabolites in the assays were annotated manually based on their (Dutch) name; using persistent IDs to annotate data is a key aspect to enable open science [37], and ultimately leads to FAIR data [38]. This annotation could not be completed for all biomarkers in the targeted assays based on their name (e.g. CysHCys, a disulfide from cysteine and homocysteine); drawing the chemical structure and converting the structure to a SMILES [39] could be used to annotate this compound. Creating pathway models (PWMs) annotated with resolvable IDs for the entities within the pathway [40] were crucial for data analysis and visualization [41]. Several initiatives merge pathway information [42,43] based on gene and protein content, rather than metabolites and chemical reactions, which makes them unsuitable for IMD metabolic data analysis. Furthermore, this reaction information is scattered over publications in images and text [44] as well as various databases [45], which requires dedicated curation time to arrive at a pathway model covering all relevant interactions. The lack of naming standardization in databases and papers for metabolic conversions cause issues in data curation, despite the IUPAC-nomenclature rules [46] and available software to translate IUPAC names to chemical structures [47] and vice versa [48]. Connecting all disease IDs to their counterpart protein ID and data from the IEMbase [29] could only be performed manually. To facilitate data integration and comparison on an automated basis, we advise providing programmatic access to biomedical databases, for example through an API [49] or SPARQL endpoint [50]. We found that some syndromes (e.g Lesch Nyhan and Kelley Seegmiller; pyrimidine 5’-nucleotidase superactivity and pyrimidine 5’-nucleotidase I deficiency) were treated as individual disorders by one database, while the other combines the information on both disorders in one entry, which hampers data interoperability. In order to distinguish between these individual disorders based on only theoretical biochemical markers, more discriminating values are needed.

### Programmers, data scientists, and bioinformaticians

By using visualization techniques from common network approaches, the developed framework is ready for extension with other data relevant for the diagnostic pipeline, e.g. genetic variants or drug-target knowledge. Furthermore, other types of (omics) data can be integrated into the workflow, for example transcriptomics, metabolomics, and fluxomics. Due to the use of semantic web technologies (RDF), other knowledge captured as Linked Open Data can also be used to extend our approach. However, this data integration requires automatable access to pathway content and open licenses; a lack herein prohibits data extraction and acquiring all relevant interactions for the studied biomarkers. Seven out of the 83 markers could not be found in any of the consulted pathway data. Several pathways overlap in terms of content, which could conceal potentially relevant pathways. Large pathways in terms of node size and captured reactions contain more biomarkers, however at a larger distance, diminishing a clear biological cause and effect path visualization. The data interpretation could be hindered when a biomarker was present only as a substrate or product, which could miss relevant up or downstream reactions. In order to overcome a mismatch of biomarkers and pathway data, clinical biomarker IDs could be converted to their corresponding neutral molecular structure InChIKey [51] ID or by performing substructure matching [52]. We want to encourage harmonizing the information in phenotype databases, for example by using existing ontologies such as the Human Disease Ontology [53], Human Phenotype Ontology [54], or Nosology for Inherited Metabolic Disorders [55]. The log2(change) data of patients was converted to the -3 to +3 scale from IEMBase, where the contribution of highly altered biomarkers to the correlation might get obscured. Furthermore, since not all clinical data could be visualized directly in one PWM (due to the biomarkers being spread over multiple models), other approaches will be needed to overcome the boundaries imposed by individual models. Reactome pathways were excluded from the analysis, since the model conversion [56] from the native Reactome pathway models to the WikiPathways RDF leads to unconnected biomarkers hampering visualization. Other possibilities to automatically visualize pathway data in the network tool Cytoscape are the Reactome Cytoscape Plugin [57] and Cytoscape for KEGG [58], the former is not optimized for metabolic data and the latter includes proprietary data access. Two other pathway apps, CyPath2 [43] and cy3sabiork [59] could not be automated.

## 4. Conclusions

With this study, we show the potential of a Systems Biology approach combining semantic web technologies for data linking and network analysis for data visualization, to directly connect biological pathway knowledge to clinical cases and biomarker data. The presented framework is adaptable to different types of IMDs, difficult patient cases, and functional assays in the future, which opens up the possibility for usage in the diagnostic pipeline. Information on treatment and clinical conditions remains important for accurate diagnosis, as well as expert interpretation of all information combined into this framework. Furthermore, several steps in the framework are now highly dependent on the manual curation of data and databases requiring expert knowledge of the information therein. The issues highlighted in the discussion section should be overcome in the future to allow our developed framework to be easily used for other IMDs, by adding persistent identifiers to (clinical) biomarker data, allowing automatable data downloads from relevant databases, and creating computer-readable pathway models from pathway figures.

## Data Availability

The data used in this study have been deposited at https://github.com/BiGCAT-UM/IMD-PUPY.

https://doi.org/10.5281/ZENODO.5632921

## Declarations

### Ethics approval and consent to participate

The Medical Research Involving Human Subjects Act (WMO) did not apply to the study mentioned above, official approval of this study by our committee was not required according to the Medical-Ethical Review committee (METC) azM/UM.

### Consent for publication

Not applicable.

### Availability of data and materials

The datasets supporting the conclusions of this article are available in the IMD-PUPY repository, https://bigcat-um.github.io/IMD-PUPY.

### Competing interests

The authors declare that they have no competing interests.

**Consortia**

The European Joint Programme on Rare Diseases (EJP RD).

## Contributions

DS: Conception and design, analysis and interpretation of data, drafted the manuscript with tables and figures (correspondence).

IH: analysis of data, performed initial review, and critically reviewed the manuscript.

CE: critically reviewed the manuscript.

JB: Conception and design, provided data, interpretation of data, performed initial review, and critically reviewed the manuscript.

EW: conducted the initial review, and critically reviewed the manuscript.

LS: conception and design, provided data, interpretation of data, performed initial review, and critically reviewed the manuscript.

## Acknowledgments

We are grateful to all participating patients and their families. Four patient samples were kindly provided by Dr. P. Fitzsimons, Dr. A. Monavari, and Dr. E. Crushell from the Children’s University Hospital and the National Centre for Inherited Metabolic Disorders, Temple Street, Ireland. We also want to acknowledge the work done by the laboratory technicians Huub Waterval, Marjon Ortmans-Ploemen, Sandra Busch, Jean-Pierre Bollen, and Karin Habets-van der Poel from the laboratory of Clinical Genetics at Maastricht University Medical Center (MUMC+).

## Abbreviations

Framework: AA: Amino acids (panel)
IDs: Identifiers
IMDs: Inherited Metabolic Disorders
Log2FC: Transformation of biomarker values to a log(2) scale
PWMs: Pathway Models
PUPY: purine and pyrimidine (panel)
RDF: Resource Description Framework
Disorders: ADA: Adenosine deaminase
ADSL: Adenylosuccinate lyase
AGXT2: Alanine--glyoxylate aminotransferase 2
AMPD1: Adenosine monophosphate deaminase 1
APRT: Adenine phosphoribosyltransferase
ARG1: Arginase 1
ASL: Argininosuccinate lyase
ASS1: Argininosuccinate synthase 1
ATIC: 5-aminoimidazole-4-carboxamide ribonucleotide formyltransferase/IMP cyclohydrolase
CPS1: Carbamoyl-phosphate synthase 1
DGUOK: Deoxyguanosine kinase
DHODH: Dihydroorotate dehydrogenase (quinone)
DPYD: Dihydropyrimidine dehydrogenase dihydropyrimidine
DPYS: Dihydropyrimidinase
HPRT1: Hypoxanthine phosphoribosyltransferase 1
IMPDH1: Inosine monophosphate dehydrogenase 1
ITPA: Inosine triphosphatase
NAGS: N-acetylglutamate synthase
NT5C3A: 5’-nucleotidase, cytosolic IIIA
OTC: Ornithine carbamoyltransferase
PNP: Purine nucleoside phosphorylase
PRPPs: (class of enzymes): Ribose-phosphate diphosphokinase
PRPS1: Phosphoribosyl pyrophosphate synthetase 1
RRM2B: Ribonucleotide reductase regulatory TP53 inducible subunit M2B
SLC25A13: Solute carrier family 25 member 13
SLC25A15: Solute carrier family 25 member 15 (ornithine translocase)
TK2: Thymidine kinase 2
TPMT: Thiopurine S-methyltransferase
TYMP: Thymidine phosphorylase
UBP1: Upstream binding protein 1
UMPS: Uridine monophosphate synthetase
XAN2: (gene: XDH): Xanthine dehydrogenase
XO: (gene: MOCOS): Molybdenum cofactor sulfurase

